# Wearable Sleep Measures May Improve Machine Learning Prediction of Home-based Pulmonary Rehabilitation Engagement Among Patients With Chronic Obstructive Pulmonary Disease: A Proof-of-Concept Study

**DOI:** 10.1101/2025.11.02.25339248

**Authors:** S. Zawada, L. Faust, M. Enayati, N. Madigan, S. Winham, R. Benzo, E. Fortune

## Abstract

**OBJECTIVE:** To evaluate whether incorporating baseline sleep measures from a wrist-worn activity monitor in machine learning (ML) models improved the prediction of 12-week engagement with home-based pulmonary rehabilitation (HBPR) in patients with chronic obstructive pulmonary disease (COPD).

**PATIENTS AND METHODS:** Among participants with a COPD exacerbation (n=124), sleep measures were collected for 1 week before HBPR and processed (1) using a validated Tudor-Locke algorithm and (2) applying partial least squares-discriminant analysis (PLS-DA) to generate the Composite Sleep Health Score. Engagement was defined as completion of one or more recommended activities per week for the 12-week duration. Nested model comparisons for logistic regression, SVM, decision tree, and naïve bayes ML models were performed to determine if including sleep measures improved engagement prediction.

**RESULTS:** In models adjusted for age, sex, Charlson Comorbidity Index, current smoker status, modified Medical Research Council score, and forced expiratory volume in 1 second, the inclusion of the Composite Sleep Health Score significantly improved the prediction of 12-week engagement only in SVM models (AUC 0.716; p=0.010). Specificity (18.2%) and accuracy (67.7%) also improved by 20.4% and 2.5%, respectively. Including the Score in the logistic regression model yielded the highest predictive performance (AUC = 0.721).

**CONCLUSION:** These proof-of-concept findings support additional investigation into the use of wearable-derived sleep measures in parametric ML models to improve screening for HBPR eligibility, identifying patients who will clinically benefit from fully remote PR. Future researchers should carefully select predictors when elucidating the link between wearable sleep measures and HBPR outcomes in COPD patients.

## BACKGROUND

Chronic obstructive pulmonary disease (COPD) is a leading cause of morbidity, disability, and mortality, ranked as the sixth leading cause of death worldwide [1]. Though established treatments, like pulmonary rehabilitation (PR), can improve COPD patient outcomes, few patients discharged after hospitalization for acute exacerbations are referred for PR participation, with some estimates finding less than 10% of eligible patients receive referrals. Complex insurance reimbursement for PR complicates eligibility screening for referrals, with many clinicians skipping the referral process due to unclear benefit over burden to patients [2]. In addition, despite PR effectiveness, attrition rates are high, with upwards of 50% of patients dropping out of PR [3]. Patients’ daily living experiences play a role in determining likelihood of study completion, with the increased transportation burden for in-person and hybrid PR contributing to low referral rates, particularly in rural areas.

Observing sleep behaviors is one strategy to understand daily living experiences of individual patients, with related measures associated with neighborhood safety, physical activity, respiratory function, and chronic disease burden [4-8]. Despite the widespread prevalence of sleep disturbances in COPD patients, the role of sleep behaviors in PR adherence and outcomes remains understudied, especially in home-based PR (HBPR) [9-11].

With the rise of precision applications in the management of respiratory diseases, the application of machine learning (ML) techniques for personalized COPD diagnosis, prognosis prediction, and treatment appears promising. One study found that baseline clinical practice data might help identify individual response to PR, suggesting a role for ML to help clinicians identify patients who would derive clinically meaningful benefits from PR [12]. Another study using baseline hospitalization data reported that ML techniques may improve 30-day rehospitalization prediction compared to traditional prediction models [13]. Despite these advances in rehospitalization and PR response prediction, most applications of ML models to acute exacerbation prediction have not demonstrated a significant improvement over conventional subjective scoring instruments. In contrast, a growing number of studies wherein ML models include wearable predictors, like accelerometer-derived steps per day, offer high prediction accuracy for exacerbations [14]. Yet, no research to date has explored the potential of wearable measures to predict PR outcomes or identify candidates most likely to complete fully remote programs, like HBPR.

Using nested model comparisons, this study evaluated the performance of multiple ML algorithms including standard COPD risk factors, with and without sleep measures derived from a wearable wristwatch at baseline, at predicting engagement throughout the duration of a 12-week home-based PR (outcome) in patients recruited after a recent COPD exacerbation at a single medical center. We hypothesized that including sleep measures with conventional PR screening criteria would improve the predictive performance of models.

## METHODS

### Datasets

Data for this study were generated by the Promoting Chronic Obstructive Pulmonary Disease Wellness through Remote Monitoring and Health Coaching clinical trial study (NCT03480386) [15]. Data on all participants and variables were extracted from the dataset collected by Mayo Clinic Division of Pulmonary and Critical Care Medicine. Electronic health record (EHR) data were retrospectively extracted for all participants with COPD in PR. Participants were recruited by an attending physician at Mayo Clinic (Rochester, MN) from 2019 to 2021. Participants had 24/7 access to health coaching staff, at no cost. Data were transmitted and stored on secure HIPAA-compliant servers.

Participants wore an activity tracker (Actigraph GT3X) on their non-dominant wrist for 7 days at baseline, prior to PR. Sleep data were processed via a Tudor-Locke algorithm optimized for chronic disease patients [16]. Participants who had contributed ≥ 3 days of sleep data were included.

Mayo Clinic institutional review board approved the study (17-009449). This study was conducted in compliance with the Declaration of Helsinki regarding ethics for medical research involving human subjects. Patients were informed of their right to object to the use of their data for the present study and gave informed consent.

### Variables

The outcome variable, engagement (yes/no), was defined as completion of one or more recommended activities (platform use, balance-related walking practice, and flexibility-related seated or standing exercises) per week for the program duration (12 weeks). Conventional predictors extracted from EHRs at baseline include the following: age, sex, Charlson Comorbidity Index (CCI), modified Medical Research Council (mMRC) score, forced expiratory volume in one second (FEV1), and current smoker status (yes/no) [16].

From the wearable data collected during the baseline period prior to HBPR, the averages of the nightly sleep quality measures were derived (Table 1).

**Table 1.** Set of optimized sleep measures derived from wearable activity tracker sensors.

| Sleep Measures (Weekly Average) |  |
| --- | --- |
| Sleep Regularity |  |
|  | Duration (minutes) |
|  | Duration (standard deviation of minutes) |
|  | Wake-after-sleep-onset (minutes) |
|  | Wake-after-sleep-onset (standard deviation of minutes) |
| Sleep Quality |  |
|  | Efficiency |
|  | Awakenings (number) |
|  | Awakenings (minutes) |
|  | Activity counts |
|  | Movement index |
|  | Fragmentation index |
|  | Sleep fragmentation index |

To account for high collinearity in wearable measures, a multivariate dimension reduction technique, partial least square discriminant analysis (PLS-DA) with 10-fold cross-validation, was applied to the wearable sleep measures. Ten-fold cross-validation was nested, ensuring that the PLS components were fitted only to training data. In PLS-DA, components are generated by maximizing splits between the outcome (yes/no). One latent variable, the first PLS component (called the “Composite Sleep Health Score” and referred to as “Score”), was included to represent wearable sleep measures in the appropriate ML models.

### Statistics

Statistical analyses were performed in R Studio 24.12.0 with the threshold significance level set at *p* < 0.05.

Descriptive statistics were summarized with percentages and frequencies by engagement status. Comparative analyses between cohorts were summarized using Mann-Whitney U (continuous) or chi-squared (categorical) tests. Considering the exploratory nature of the study and its small sample size n (< 1000), model performance was evaluated using 10-fold cross-validation, in which the dataset is randomly split into 9 training and 1 test sets, or folds. The ML model is trained on the 9 training folds and the 1 test fold is used for independent validation. This is performed 10 times, with the results from each fold averaged, using the R software package *caret*. The trainControl function of the R package *caret* applies cross-validation using automated random sampling. To train models, the *train* function of the *caret* package was used, offering automatic parameter tuning, selection of optimal models, and performance estimates.

To assess the extent to which 12-week engagement in PR can be automatically discriminated from dropouts based on standard clinical data and whether the inclusion of baseline sleep measures can enhance the predictive performance of such models, four ML techniques were used: (1) logistic regression classification, with a generalized linear model and logit link function; (2) decision tree classifier, built with R software package *rpart*, used classification and regression tree (CART) to assemble binary classification trees; (3) support vector machine (SVM) model built using the R software package *e1071*; and (4) Naïve Bayes decision support model built using the R software package *klaR* [16].

To summarize performance, accuracy, sensitivity, and specificity were averaged across folds for each model. Accuracy was defined as the proportion of true results, whether true-negatives or true-positives, classified correctly by a model. Specificity and sensitivity were defined as the proportion of true-negatives and true-positives classified correctly, respectively. After the receiver operator characteristic (ROC) curve for a model was graphed, the area under the ROC curve (AUC) was calculated. For each ML technique, a nested comparison of performance (AUC) with and without wearable sleep measures was performed using DeLong’s test. For each model, importance plots were charted to graphically compare the weights of predictors in each algorithm.

### Sensitivity Analysis

To assess the contribution of the Score to model prediction performance, the first sensitivity analysis assessed the model with the Score only, excluding other predictors. A second sensitivity analysis dropped the Score and included sleep regularity measures: mean sleep duration, mean WASO, standard deviation of sleep duration, and standard deviation of WASO. Using direct measures in lieu of the Score facilitated comparison between the predictive power of original measures of sleep and the variance maximizing predictor (the Score).

## RESULTS

The baseline characteristics of the cohort by engagement status are presented in Table 2. The distribution of mMRC values in the cohorts were different. The proportion of current smokers was higher in the dropout cohort.

**Table 2.** Comparison of characteristics of the engaged and dropout cohorts of the dataset.

|  | Remained Engaged for the 12-Week HBPR |  | p-value |
| --- | --- | --- | --- |
|  | Yes (n = 80) | No (n = 44) |  |
| Male (% , n) | 52.5% (42) | 36.4% (16) | 0.125 |
| Age (Median [IQR]) | 71 [64-78] | 67 [62-73] | 0.086 |
| CCI (Mean ± SD) | 5 (3.4) | 5 (2.8) | 0.859 |
| FEV1 (Mean ± SD) | 47.9 (20.6) | 42.8 (17.9) | 0.185 |
| Current Smoker (% , n) | 3.8% (3) | 18.1% (8) | **0.018** |
| College Degree (% , n) | 25% (20) | 20.5% (9) | 0.726 |
| mMRC (Mean ( SD)) | 2 (0.9) | 3 (0.7) | **0.001** |
| WASO Duration (Mean (SD)) | 74.8 (35.8) | 80.9 (45.8) | 0.569 |
| Sleep Duration (Mean (SD)) | 526.0 (115.0) | 493.7 (108.5) | 0.182 |

In terms of predicting which participants with COPD would remain engaged over the 12-week HBPR, the highest model performance was obtained by logistic regression classification when the Score was included (AUC = 0.721), followed by the AUC from SVM classification with the Score (AUC = 0.716; Figure 1). The lowest model performance was obtained by decision tree without the Score (AUC = 0.548). The inclusion of the Score in the SVM model significantly improved its performance (*p =* 0.010) (Figure 1), but no statistically significant findings were observed in other nested model comparisons.

**Figure 1.**
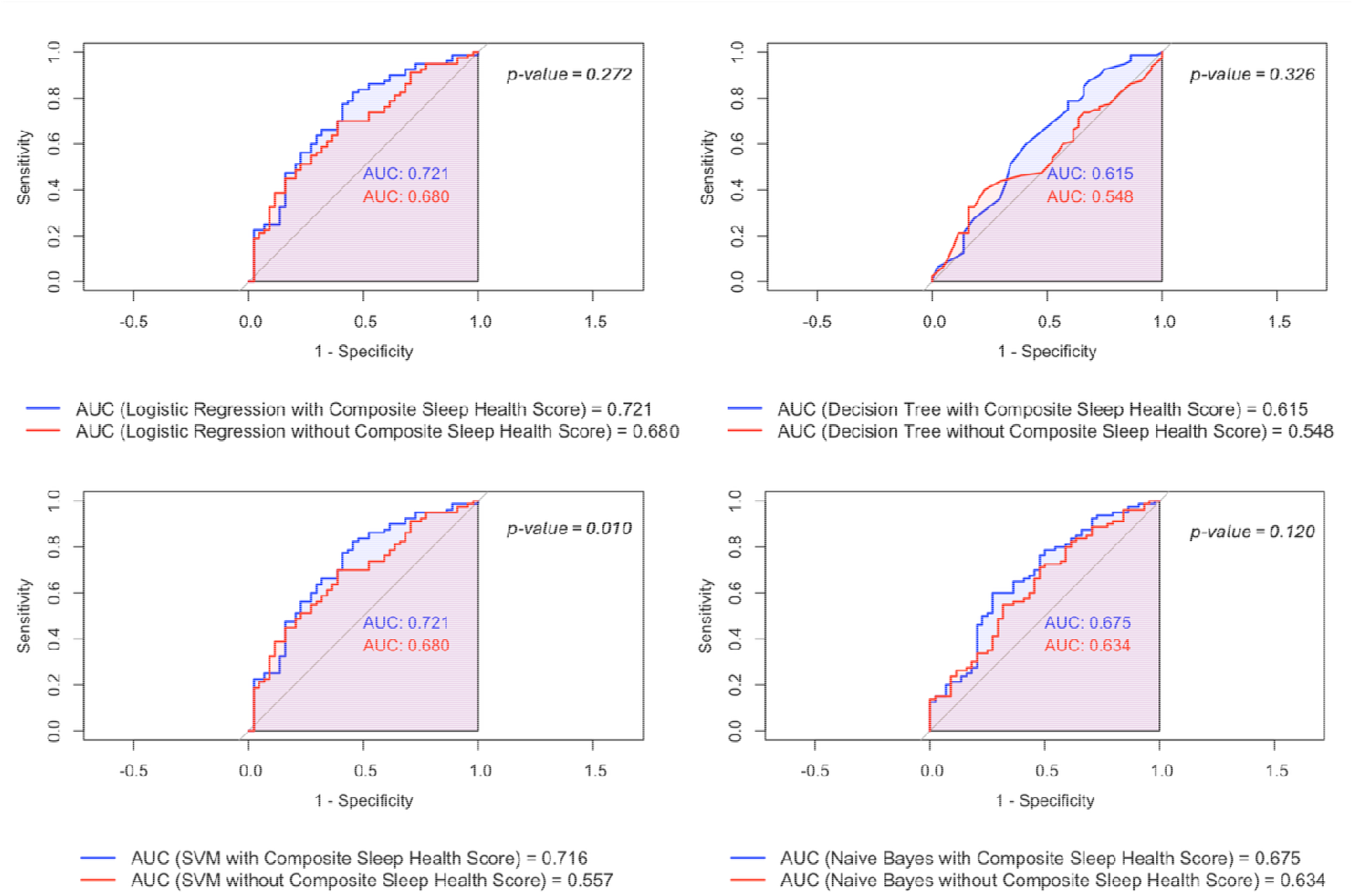
Comparisons of AUC Curves (Nested Model Comparisons)

In the logistic regression model, the Score improved the accuracy (+4.9%) and specificity (+18.2) with a slight reduction in sensitivity (-2.4%) (Table 3). In contrast, inclusion of the Score in the decision tree model resulted in no change in specificity, while increasing sensitivity (+11.2%) and accuracy (+7.2). Adding the Score to the Naïve Bayes model did not change sensitivity, but improved specificity (+9.0%) and accuracy (+3.2%). The SVM model with Score showed improvement in specificity (+20.4%) and accuracy (+2.5%) with a reduction in sensitivity (+7.5).

**Table 3.**
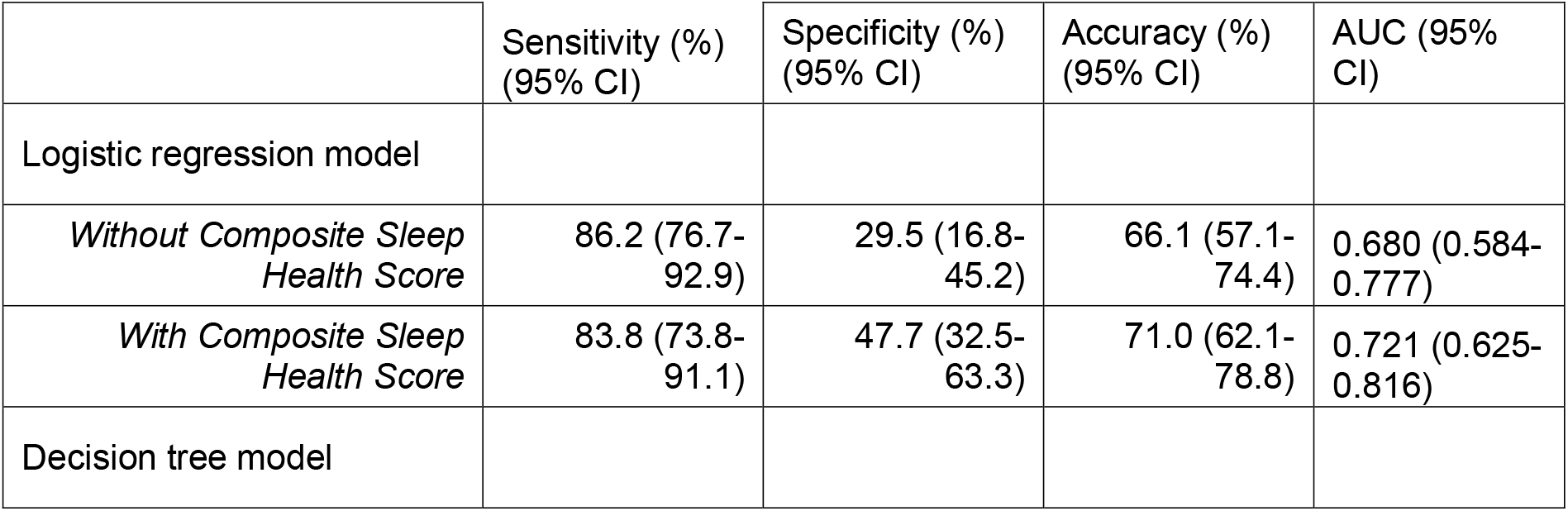
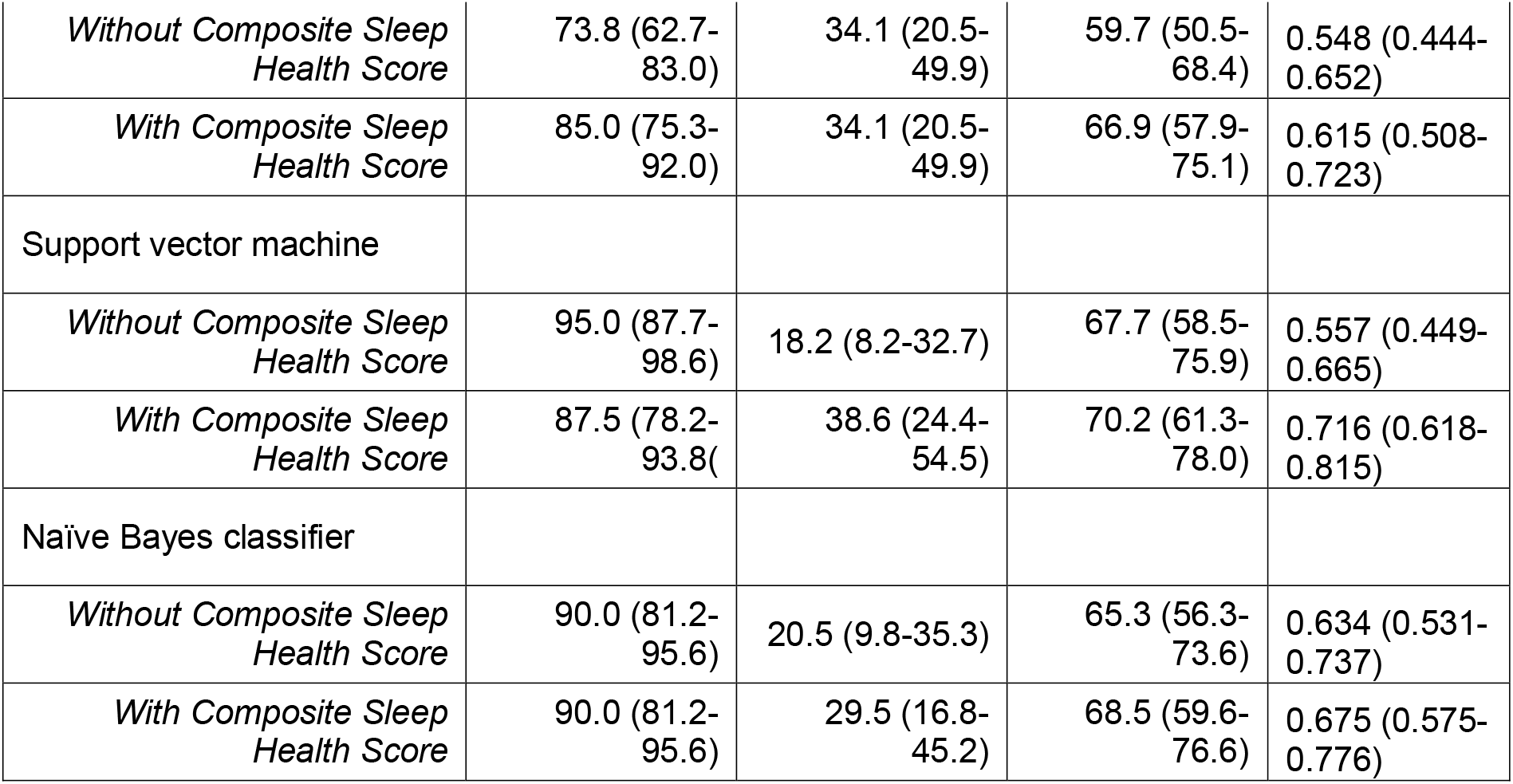
Prediction of 12-week study engagement in HBPR patients with COPD using ML techniques.

|  | Sensitivity (%)<br>(95% CI) | Specificity (%)<br>(95% CI) | Accuracy (%)<br>(95% CI) | AUC (95%<br>CI) |
| --- | --- | --- | --- | --- |
| Logistic regression model |  |  |  |  |
| *Without Composite Sleep Health Score* | 86.2 (76.7-92.9) | 29.5 (16.8-45.2) | 66.1 (57.1-74.4) | 0.680 (0.584-0.777) |
| *With Composite Sleep Health Score* | 83.8 (73.8-91.1) | 47.7 (32.5-63.3) | 71.0 (62.1-78.8) | 0.721 (0.625-0.816) |
| Decision tree model |  |  |  |  |
| *Without Composite Sleep Health Score* | 73.8 (62.7-83.0) | 34.1 (20.5-49.9) | 59.7 (50.5-68.4) | 0.548 (0.444-0.652) |
| *With Composite Sleep Health Score* | 85.0 (75.3-92.0) | 34.1 (20.5-49.9) | 66.9 (57.9-75.1) | 0.615 (0.508-0.723) |
| Support vector machine |  |  |  |  |
| *Without Composite Sleep Health Score* | 95.0 (87.7-98.6) | 18.2 (8.2-32.7) | 67.7 (58.5-75.9) | 0.557 (0.449-0.665) |
| *With Composite Sleep Health Score* | 87.5 (78.2-93.8) | 38.6 (24.4-54.5) | 70.2 (61.3-78.0) | 0.716 (0.618-0.815) |
| Naïve Bayes classifier |  |  |  |  |
| *Without Composite Sleep Health Score* | 90.0 (81.2-95.6) | 20.5 (9.8-35.3) | 65.3 (56.3-73.6) | 0.634 (0.531-0.737) |
| *With Composite Sleep Health Score* | 90.0 (81.2-95.6) | 29.5 (16.8-45.2) | 68.5 (59.6-76.6) | 0.675 (0.575-0.776) |

Importance plots visualizing the weights of variables for models predicting 12-week study engagement with predictor weights are in Figure 2. In all models without the Score, mMRC scores had the highest importance ranking. In SVM and logistic regression models, including the Score dropped mMRC importance ranking by one level, with the Score as the variable of highest importance. Including the Score in the decision tree model resulted in a reordering variable importance, with importance rankings for CCI score, FEV1, and current smoker status higher than for mMRC. In the Naïve Bayes model with the Score, the Score was the variable of least importance.

**Figure 2.**
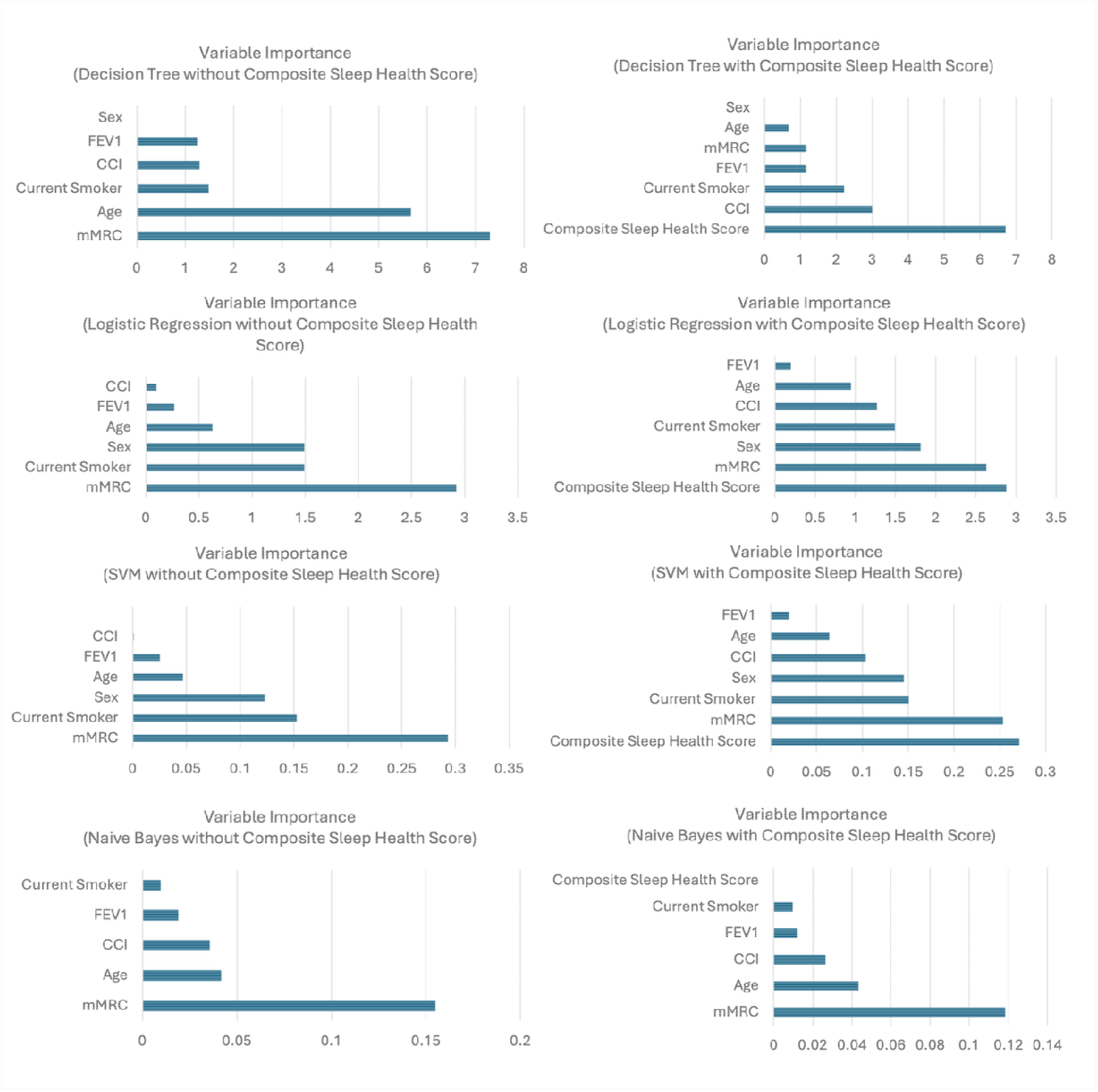
Variable Importance Graphs

In the first sensitivity analysis, using only the Score with no conventional predictors, the significant SVM model performance improvement was lost (Supplementary Table 1). For most models with the Score alone, predictive performance decreased. The logistic regression classification model without the Score generated the highest AUC curve (0.708).

Similar results were generated in the second sensitivity analysis (Supplementary Table 2). Although an improvement in predictive performance was observed in the SVM model when sleep quality measures were included with conventional predictors, this was not statistically significant.

## DISCUSSION

In this exploratory study, we compared ML model performance with and without sensor-derived sleep measures from baseline for the prediction of 12-week engagement with HBPR in a cohort of patients with COPD. When assessing if the addition of the Score offered an advantage for predictive performance, only one ML technique (SVM) yielded statistically significant improvement; however, the logistic regression model with the Score generated the highest predictive performance. Considering the sensitivity analyses, with the drop in performance for the SVM model using the Score as the sole predictor or sleep quality measures as predictors in addition to conventional predictors, baseline sleep measures without dimensionality reduction processing and conventional predictors may not be useful for predicting 12-week HBPR engagement; however, when the Score is included with conventional predictors used for COPD PR eligibility assessment, like mMRC, CCI, and FEV1, this approach could have applications in HBPR eligibility screening. For instance, patients with low prediction scores for 12-week HBPR engagement might be more appropriately referred to hybrid or in-person PR.

Although the inclusion of the Score in models did not offer outstanding predictive value for 12-week HBPR engagement, it did yield moderate discriminatory performance beyond chance in the SVM and logistic regression models. Despite the relatively low AUC for the SVM model without the Score, the SVM model with the Score yielded the second highest predictive performance overall, after the logistic regression model with the Score. For other models, inclusion of the Score with conventional predictors improved predictive performance beyond that of only conventional predictors, though not statistically significant. Combined, these results suggest parametric ML models may be appropriately suited to using wearable sleep data to predict HBPR outcomes in future research. Since parametric techniques are more resistant to overfitting to noise, it is plausible that the relationship between the predictors and outcome may be subtle.

Considering the low-to-moderate predictive performance in all models (AUC < 0.800), and the high predictive performance observed in the models using parametric techniques, the relationship between sleep and HBPR engagement status might be straightforward and, therefore, not benefit from the application of ML techniques. ML is effective at identifying differences in large datasets with complicated relationships between predictors and outcomes but is less useful in less complex datasets. For instance, Moll et al applied a multimodal approach to predicting mortality in COPD patient cohorts using baseline physical activity (6-minute walk distance, 6MWT), mMRC, and FEV1 values – as well as an imaging predictor (pulmonary artery-to-aorta ratio) [20]. With their large dataset (n = 2,632), the performance of ML models was similar to that of traditional statistical methods; however, the baseline 6MWT was performed in clinical settings, unlike our collection in real-world settings. Thus, the use of passively collected data during the eligibility screening phase could be useful for patients unable to visit for conventional in-person assessments used for PR eligibility screening, thereby reducing barriers to HBPR.

Longer longitudinal data collection might be necessary to improve ML models using wearable measures. A study by Wu et al including a wearable sleep metric and home air-quality sensor measures in real-world settings generated a 7-day exacerbation prediction model using a deep neural network technique with high sensitivity (94%), specificity (90.4%), accuracy (92.1%), and AUC values (>0.900). Although the cohort size was small (n = 67), this longitudinal study collected data for 16 weeks, far beyond the 1-week study design common in the wearables literature, and further supporting the rationale that ML classification algorithms perform better with multimodal, longitudinal datasets when only smaller sample sizes are available [25]. The interpretation of our results is limited by the fact that most research in the COPD space to date has been applied to exacerbation and readmission prediction, not PR engagement. Additionally, little research into fully remote PR has been conducted [26]. In contrast, multiple studies applying ML techniques to predict response to in-hospital PR have been published. For instance, Vitacca et al. investigated the probability of success in in-hospital PR, applying clustering analysis. They found that the cluster with the most severe profile achieved the highest percentage of clinically meaningful PR improvements; however, this study used only in-person baseline data [27]. A growing number of studies assessing sleep in PR suggest that completing a PR program is associated with improved sleep health; however, the directionality of the relationship, if any, between PR and sleep is unknown [28-29].

The chief limitation of this study is its small sample size, which limited the set of ML techniques used [30]. For instance, none of the ML techniques consider spatial relationships. The low predictive values observed in this study might also be explained by the small sample size. Current research suggests that cross-validation yields biased estimates in sample sizes less than 1000 [31]. Considering that ML algorithm bias can be mitigated by increasing sample size, our sample (n= 124) may have introduced variance in the prediction of classification. Another limitation is that the outcome, 12-weeks of engagement, was designed as a binary classifier. As prior evidence suggests many COPD patients, regardless of COPD severity, benefit, either physically or psychologically from PR, future research could consider HBPR engagement as a continuous variable, applying ML regression techniques instead of classification ML techniques [32]. An additional limitation is that participants with sleep-related conditions that are highly prevalent in COPD populations, like obstructive sleep apnea, were not excluded from in this study. Since sleep disturbance symptoms emerge over time, and are often overlooked, it is likely that some patients in the cohort have undiagnosed sleep conditions. Moreover, for those with pre-existing sleep diagnoses, management of sleep conditions is overwhelmingly poor [33]. Rather than considering sleep diagnoses exclusively, we opted to assess sleep behavior regardless of sleep disorder diagnoses [34]. Our study is also limited by only including internal validation with 10-fold cross-validation. As our study lacked external validation, with no models tested on other comparable datasets from HBPR participants, the reproducibility and generalizability of the models examined in this study remain unclear.

Overall, these results highlight the potential of parametric ML techniques to capture subtle effects in wearable sleep measures for HBPR engagement status classification. Though no identical study is available for comparison, our significant findings suggest that SVM may be the preferred ML technique for predicting HBPR engagement with baseline sleep data in COPD cohorts, a finding consistent with COPD literature [17]. SVM regularly outperforms other ML techniques when dealing with high-dimensional data, like that from wearable sensors, and small sample sizes [18-19]. Future work should consider additional modes of data, from blood biomarker to audio recordings, as possibly key to unlocking the potential of parametric ML techniques applied to HBPR engagement prediction beyond the slight improvements in model performance observed here [21-24]. Also beyond the scope of this study, the improved performance observed when the Score was included in the SVM models could be attributable to unknown COPD subtypes, detected via sleep metrics, which might elucidate engagement trends in PR. No research, to our knowledge, has explored sleep health interventions in HBPR or a clinically meaningful difference in sleep regularity during PR. With this exploratory study as a starting point for researchers to generate hypotheses about sleep in HBPR, using multimodal datasets with wearable measures is ripe for discovering new insights into engagement and improving attrition.

## CONCLUSION

Efficient eligibility screening for HBPR participation is critical to addressing high attrition rates. Although the inclusion of wearable sleep measures, captured as the Score, in the SVM model significantly improved the predictive performance of 12-week HBPR engagement in patients with COPD, the logistic regression model with the Score exhibited the highest predictive performance. The results from parametric ML models may help inform treatment decisions for COPD, such as recommending HBPR over in-hospital PR based on the likelihood of remaining engaged over the 12-week remote study period. Future research should prioritize larger cohort enrollment and longer longitudinal study designs as well as a thoughtful approach to the inclusion of multimodal data, such as imaging, genetic, and respiratory rate predictors to further validate the use of wearable sleep measures in HBPR research.

## Data Availability

All data produced in the present study are available upon reasonable request to the authors

## ABBREVIATIONS

ML: machine learning
COPD: chronic obstructive pulmonary disease
PR: pulmonary rehabilitation
HBPR: home-based pulmonary rehabilitation
PLS-DA: partial least squares-discriminant analysis
SVM: support vector machine
CCI: Charlson comorbidity index
mMRC: modified medical research council
FEV1: forced expiratory volume in 1 second
AUC: area under ROC curve
ROC: receiver operating characteristic

## Ethics Statement

The institutional review board at Mayo Clinic approved the study (17-009449). This study was conducted in compliance with the Declaration of Helsinki regarding ethics principles for medical research involving human subjects. Patients were informed of their right to object to the use of their data for the present study and gave their informed consent.

## Notes

**Conflict of interest disclosure:** The authors report no conflicts of interest.

### Competing Interest Statement

The authors have declared no competing interest.

### Funding Statement

Funding for this study was provided by NIH R56 HL173214, NIH R01 HL140486, and the Robert D. and Patricia E. Kern Center for the Science of Health Delivery. Funding for the Prostate Cancer and AI Falls Projects is from the Robert D. and Patricia E. Kern Center for the Science of Health Delivery.

