## Supplementary Tables 1 and 2 for "Wearable Sleep Measures May Improve Machine Learning Prediction of Home-based Pulmonary Rehabilitation Engagement Among Patients With Chronic Obstructive Pulmonary Disease: A Proof-of-Concept Study"

**Supplementary Material**

SENSITIVITY ANALYSIS #1 (Composite Sleep Health Score only with no other predictors)

|  | Sensitivity (%) | Specificity (%) (95% CI) | Accuracy (%) (95% CI) | AUC (95% CI) | p-value for AUCs |
| --- | --- | --- | --- | --- | --- |
| Logistic regression model |  |  |  |  | 0.119 |
| *Without Composite Sleep Health Score* | 85.0 (75.3-92.0) | 31.8 (18.6-47.6) | 66.1 (57.1-74.4) | 0.708 (0.611-0.805) |  |
| *With Composite Sleep Health Score* | 91.2 (82.8-96.4) | 25.0 (13.2-40.3) | 67.7 (58.8-75.9) | 0.595 (0.484-0.706) |  |
| Decision tree model |  |  |  |  | 0.917 |
| *Without Composite Sleep Health Score* | 87.5 (78.2-93.8) | 36.4 (22.4-52.2) | 69.4 (60.4-77.3) | 0.511 (0.403-0.619) |  |
| *With Composite Sleep Health Score* | 88.8 (79.7-94.7) | 22.7 (11.5-37.8) | 65.3 (56.3-73.6) | 0.520 (0.405-0.635) |  |
| Support vector machine |  |  |  |  | 0.654 |
| *Without Composite Sleep Health Score* | 96.2 (89.4-99.2) | 18.2 (9.2-32.7) | 68.5 (59.6-76.6) | 0.593 (0.485-0.701) |  |
| *With Composite Sleep Health Score* | 93.8 (86.0-97.9) | 13.6 (5.2-27.4) | 65.3 (56.3-73.6) | 0.559 (0.450-0.669) |  |
| Naïve Bayes classifier |  |  |  |  | 0.087 |
| *Without Composite Sleep Health Score* | 97.5 (91.3-99.7) | 22.7 (11.5-37.8) | 71.0 (62.1-78.8) | 0.680 (0.611-0.805) |  |
| *With Composite Sleep Health Score* | 88.8 (79.7-94.7) | 22.7 (11.5-37.8) | 65.3 (56.3-73.6) | 0.548 (0.484-0.706) |  |

SENSITIVITY ANALYSIS #2 (No Composite Sleep Health Score, replaced with sleep regularity direct measures)

|  | Sensitivity (%) | Specificity (%) (95% CI) | Accuracy (%) (95% CI) | AUC (95% CI) | p-value for AUCs |
| --- | --- | --- | --- | --- | --- |
| Logistic regression model |  |  |  |  | 0.306 |
| *Without Composite Sleep Health Score* | 87.5 (78.2-93.8) | 31.8 (18.6-47.6) | 67.7 (58.8-75.9) | 0.695 (0.599-0.791) |  |
| *With Composite Sleep Health Score* | 86.2 (76.7-92.9) | 27.3 (15.0-42.8) | 65.3 (56.3-73.6) | 0.627 (0.517-0.737) |  |
| Decision tree model |  |  |  |  | 0.737 |
| *Without Composite Sleep Health Score* | 73.8 (62.7-83.0) | 40.9 (26.3-56.8) | 62.1 (52.9-70.7) | 0.533 (0.424-0.641) |  |
| *With Composite Sleep Health Score* | 71.2 (60.0-80.8) | 43.2 (28.3-59.0) | 61.3 (52.1-69.9) | 0.560 (0.451-0.669) |  |
| Support vector machine |  |  |  |  | 0.684 |
| *Without Composite Sleep Health Score* | 93.8 (86.0-97.9) | 22.7 (11.5-37.8) | 68.5 (59.6-76.6) | 0.566 (0.456-0.675) |  |
| *With Composite Sleep Health Score* | 92.5 (84.4-97.2) | 18.2 (8.2-32.7) | 66.1 (57.1-74.4) | 0.595 (0.489-0.702) |  |
| Naïve Bayes classifier |  |  |  |  | 0.082 |
| *Without Composite Sleep Health Score* | 95.0 (87.7-98.6) | 18.2 (8.2-32.7) | 67.6 (58.8-75.9) | 0.679 (0.599-0.791) |  |
| *With Composite Sleep Health Score* | 82.5 (72.4-90.1) | 22.7 (11.5-37.8) | 61.3 (52.1-69.9) | 0.554 (0.517-0.737) |  |
